# Salt and Health: Public awareness, attitudes and practices in Sri Lanka to inform a behavior change communication campaign to reduce dietary salt

**DOI:** 10.1101/2020.07.25.20162081

**Authors:** Achala Jayatilleke, Nalika Gunawardena, Angela de Silva, Champika Wickramasinghe, Lakshman Gamlath, Thilak Siriwardena, Vindya Kumarapeli, Janaki Vidanapathirana, Shanthi Gunawardena, AMAAP Alagiyawanna, Ishanka Talagala, Aravinda Wickramasinghe, Prabha Kumari, Prasad Ranatunga, Sapumal Dhanapala, Razia Pendse

**Affiliations:** World Health Organization (WHO) Country Office for Sri Lanka, Colombo, Sri Lanka; World Health Organization (WHO) Regional Office of South East Asia, New Delhi, India; Ministry of Health and Indigenous Medical Services, Colombo, Sri Lanka

## Abstract

**Background:** Sri Lankan citizens consume almost double the recommended daily amount of salt. Objective of this study was to assess the knowledge, attitudes and practices related to health effects of dietary salt among adults and adolescents in Sri Lanka to inform a national behavior change communication campaign.

**Methods:** We conducted a descriptive household survey among adults (n=1016) and adolescents (n=505) in 10 districts. An, interviewer administered questionnaire was used for data collection. The approximate amount of dietary salt intake of the individuals was estimated based on household purchases.

**Findings:** The recommended salt limit was identified by 40% of the population. Majority, adults (90.8%) and adolescents (86.1%) knew the adverse health effects of high salt intake. Although household monthly purchase of salt indicated consumption is much higher than recommended, 48.3% of adults and 45.9% of adolescents believed that they consume ‘just the right’ amount. Discretionary salt added to home cooking was a major contributor to intake, with approximately half (50%) adding salt when cooking rice, the staple. For health-related information most preferred (adults - 72%, adolescents – 69%) media is television.

**Interpretation:** The study identified gaps as well strengths in knowledge, attitudes and practices of Sri Lankans related to salt and health and recommends that the communication campaign include specific messaging to address gaps and leveraging on strengths. The survey identified adult females to be a key target group for the campaign and television is recommended as the mode of delivery.

## Introduction

Convincing evidence suggest that high salt intake is associated with an increased risk of hypertension and in turn, coronary artery disease, and stroke^1^. Furthermore, a review on salt and health and current experience of worldwide salt reduction programmes in 2012 revealed that beyond the effect of salt intake on blood pressure, high salt intake may directly increase the risk of strokes, left ventricular hypertrophy, osteoporosis, stomach cancers and renal diseases^2,3^.

Reducing salt in food as a measure to reduce population level consumption of salt has been identified as a cost-effective intervention to prevent non-communicable diseases by the World Health Organization (WHO). It recommends that adults should consume less than 5g of salt per day^4^.

A review of evidence of salt intake of different countries in 2019 to estimate the global salt intake revealed that salt intake is higher than the WHO recommended level of 5g of salt per day^5^. In most high-income countries, bulk of dietary salt (70-80%) come from processed or restaurant foods^1^. Processed foods such as breads, processed meat and snack foods, as well as condiments (e.g. soy sauce, fish sauce) have been found to be the source of dietary salt in high income countries. Evidence on the source of dietary salt in low- and middle-income countries is scarce.

Sri Lanka is an upper-middle income country in the South East Asia region of the WHO with a total population of approximately 22 million. All evidence indicates that per capita salt intake to be much higher than the WHO recommended level. National Population Salt Consumption survey in 2012 showed that estimated salt intake of a person to be 10.5g/day, double the recommended amount of salt intake^6^. A recent study reported in 2020 conducted among 328 adults between 30-59 years resident in one of the nine provinces in the country using the 24-hour urinary excretion, revealed a mean daily salt consumption of 8.3g/day^7^.

Evidence on sources of dietary salt indicate that the bulk of dietary salt in the country comes from discretionary salt added to home cooked foods. The 2015 STEPs survey in Sri Lanka conducted among nationally representative sample of 5188 adults aged 18-69 years assessed various sources of dietary salt in Sri Lankan households. The survey revealed that adding salt to rice, the staple food while cooking could be one important sources of dietary salt with more than half of the target households (52.8%) reported adding salt to rice^8^. With regards to consuming processed foods high in salt (including informal processed food like ‘*wade’*, etc.), approximately 27% of the adults (28.3% males and 24.8% females) were estimated to always or often eat such processed food. However, the STEPS survey didn’t provide adequate information on knowledge, attitudes and practices of salt consumption to organize a campaign for behavioral change^8^.

Sri Lanka, committed to implement the National Salt Reduction Strategy 2018-2022 (NSRS) with the overall aim of a 30% reduction in mean population intake of salt/sodium by 2025 from its baseline of 10.5g/day in 2012, which was determined by a national survey^6,9^. The main justification for NSRS was to mitigate the country’s’ high burden of hypertension. Prevalence of hypertension among adults was 26.1% (males 25.4%; females 26.7%) in 2015 according to the STEPs survey^8^. Guided by the SHAKE technical package of WHO^4^, the NSRS calls for better surveillance, implementation of an effective behavior change communication and mass media campaign, harnessing food industry to reformulate food products to contain less salt, adopting a front of pack labeling system and establishment of a supportive environment in the community for reduced salt consumption. Given the available evidence on the sources of dietary salt in Sri Lanka, designing and implementing an effective behavior change communication and mass media campaign is considered to be an imperative strategy to be prioritized in reaching the intended aim of salt reduction in the country. Designing communication materials for behavior change of communities, requires specific information on salt containing food consumption of individuals, their awareness on the adverse effects of high salt consumption, and their attitudes reflecting the readiness for change. The objective of the present national level survey was to assess the knowledge, attitude and practices of adolescents aged 15-18 years and adults related to salt and health, to inform the designing of communication material in implementing an effective behavior change communication campaign for the NSRS 2018-2022.

## Methods

### Study design and duration

We conducted an island wide cross-sectional descriptive household survey during August and September 2019, in 10 of the 25 districts of Sri Lanka.

### Study population

The study population included two population age groups. Adult males and females aged 19 to 69 years and adolescents aged 15-18 years who had been resident in the district for more than six months. We excluded those mentally disturbed and physically too frail to participate in the study.

### Sample size

We calculated the minimum required numbers of adults and adolescents for this survey based on the number required to estimate the proportion of study units who can be expected to have good knowledge, attitudes and practices related to salt and health, using the formula “N=z^2^×P×(1−P)/e^2^” (z=level of confidence (for a level of confidence of 95%, z=1.96); P=estimated proportion of the population that presents the characteristic, P=0.5; e=margin of error, e=0.05. With z being 1.96, the minimum required sample size was 384 participants^10^. To account for the design effect due to the adopted multistage cluster sampling technique in the household survey and possible non-responses, the minimum required sample size was considered as 500. Considering the importance of capturing the variation of knowledge, attitudes and practices, we included 500 each from each category of adult males, adult females, and adolescents with a total, of 1500 participants.

### Sampling method

We used a multi-stage cluster sampling technique to select a representative sample of adults and adolescents from 10 districts of Sri Lanka with numbers from each district being proportionate to size of the resident population. Sri Lanka has nine provinces. In the first stage, we randomly selected one district from each province and in addition to the nine selected, the Colombo district, which is the most populous district of Sri Lanka; was also included making the total number of districts ten out of the 25 districts in the country.

We defined a cluster as a group of 54 eligible study units - 36 adults (18 males and 18 females) and 18 adolescents living in a basic medical administrative area which has around 100,000 population. (Medical officer of Health (MOH) area) We required 28 such clusters to fulfill the sample of 1500 and these 28 clusters were assigned to the 10 districts proportionate to the population in the district. The required number of MOH area clusters were then selected randomly from a sampling framework MOH areas of each of the district.

The eligible study units residing in households located in a randomly picked a road within the MOH area were invited to participate in the study by visiting the consecutively located households until the required number of study units of all three categories were recruited. We ensured that only one study unit from one of the three categories of adult males, adult females or adolescents was recruited from one household and used the Kish method to select a member from the house for the survey^11^.

### Study instrument

Based on a desk review of published literature on surveys with similar objectives^12–18^, we developed an interviewer administered multicomponent knowledge, attitude, and practice survey questionnaire on salt and health as an online version using EpiCollect5 free data gathering platform^19^. in the present study, we estimated the approximate amount of per capita dietary salt of the households of adults included in the study by inquiring into the amount of monthly purchases of salt to the households and the number of household members. Considering the fact that this survey was conducted to inform the design of the communication package to support the implementation of NSRS 2018-2022, specific inquiries were also made on the commonly used information source to access health information.

### Data collection

We trained and used a team of 28 undergraduate university students studying in the “Health Promotion” stream to collect data in the household survey, assigning one data collector for each cluster (MOH area). Data were collected using smart phones and were uploaded to be in real time with the geolocation to the server. We used geolocations and time stamps to remotely monitor the data collection process.

### Data analysis

We report descriptive analysis of the knowledge, attitude and practices of adolescents aged 15-18 years and adults, regarding salt and health using frequencies distributions and used Chi-square test to assess for any gender differences in the results.

### Ethical considerations

We obtained ethical approval for this study from the Ethics Review Committee of the Postgraduate Institute of Medicine, University of Colombo (PGIM/ERC/2019-140). Participation in this study was voluntary and we obtained informed written consent from all the participants prior to the study.

## Results

As illustrated in Table 1, 1016 adults participated in this study. The median age of the adult participants was 46 years with an inter quartile range (IQR) of 35-58 years. Information related to years of formal education showed that approximately three fourths of adults (n=742; 76.3%) (males-n=373; 76.0%: female-n=369; 76.5%) had over 10 years of education. Median monthly household income of adult participants was Rs. 34,000 (IQR: Rs. 20,000-Rs. 50,000). Among adults, 26.7% (n=270) (males-n=129; 25.4%: females-n=141; 27.9%) were having hypertension. This is compatible with the findings of the 2015 STEPS survey that reported 26.1% of the adults had raised blood pressure^8^. Moreover, 7.6% (n=77) (males-n=49; 9.7%: female-n=28; 5.6%) had heart diseases and 2.0% (n=20) (males-n=16; 3.2%: female-n=4; 0.8%) reported kidney diseases. Television (TV) was by far the most commonly used information source to access health information among adults (762; 76.6%). However, the proportion of adult females (n=404; 81.3%) who accessed TV for health information was significantly higher than the adult males (n=358; 71.9%) (p=0.008).

**Table 1.**
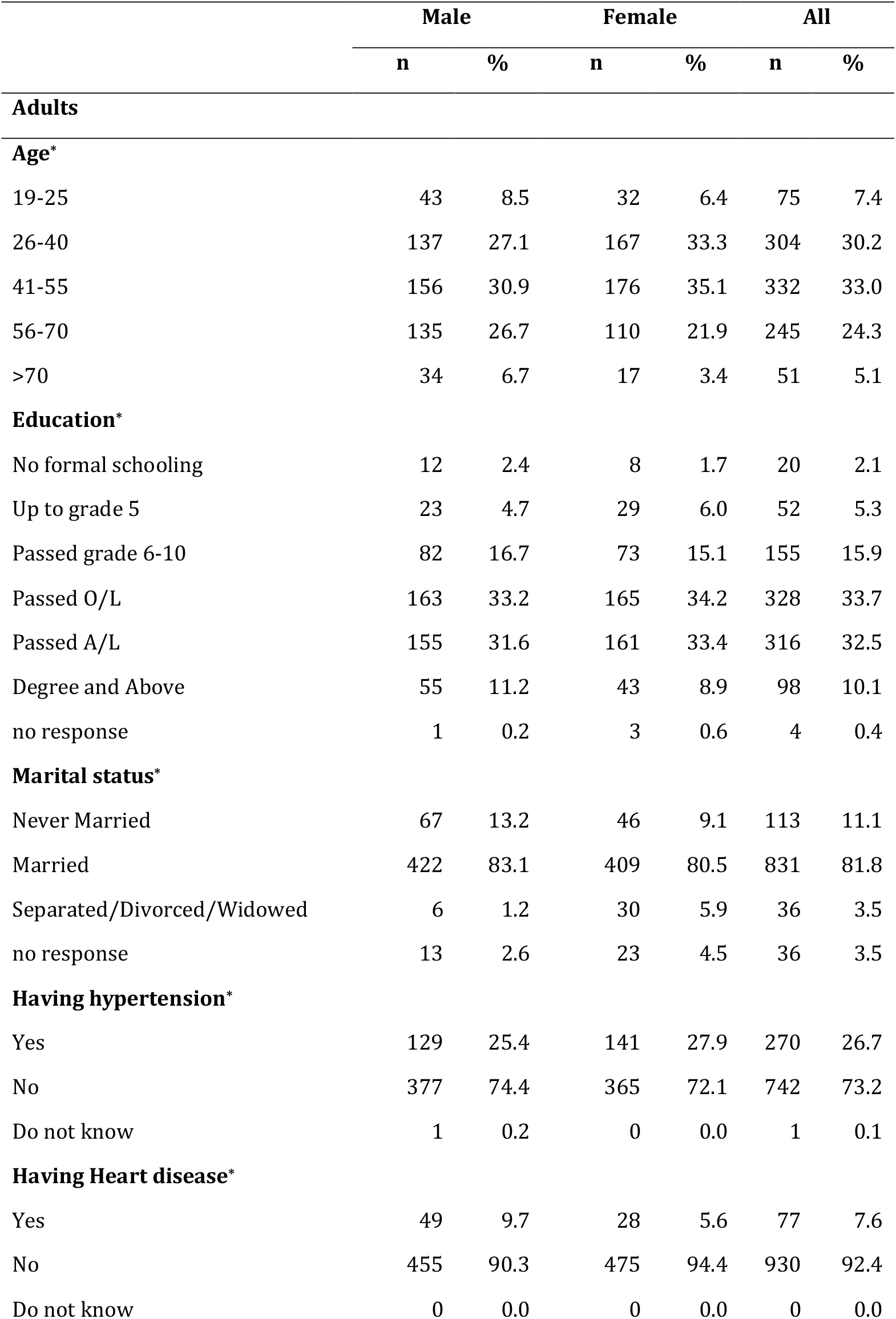
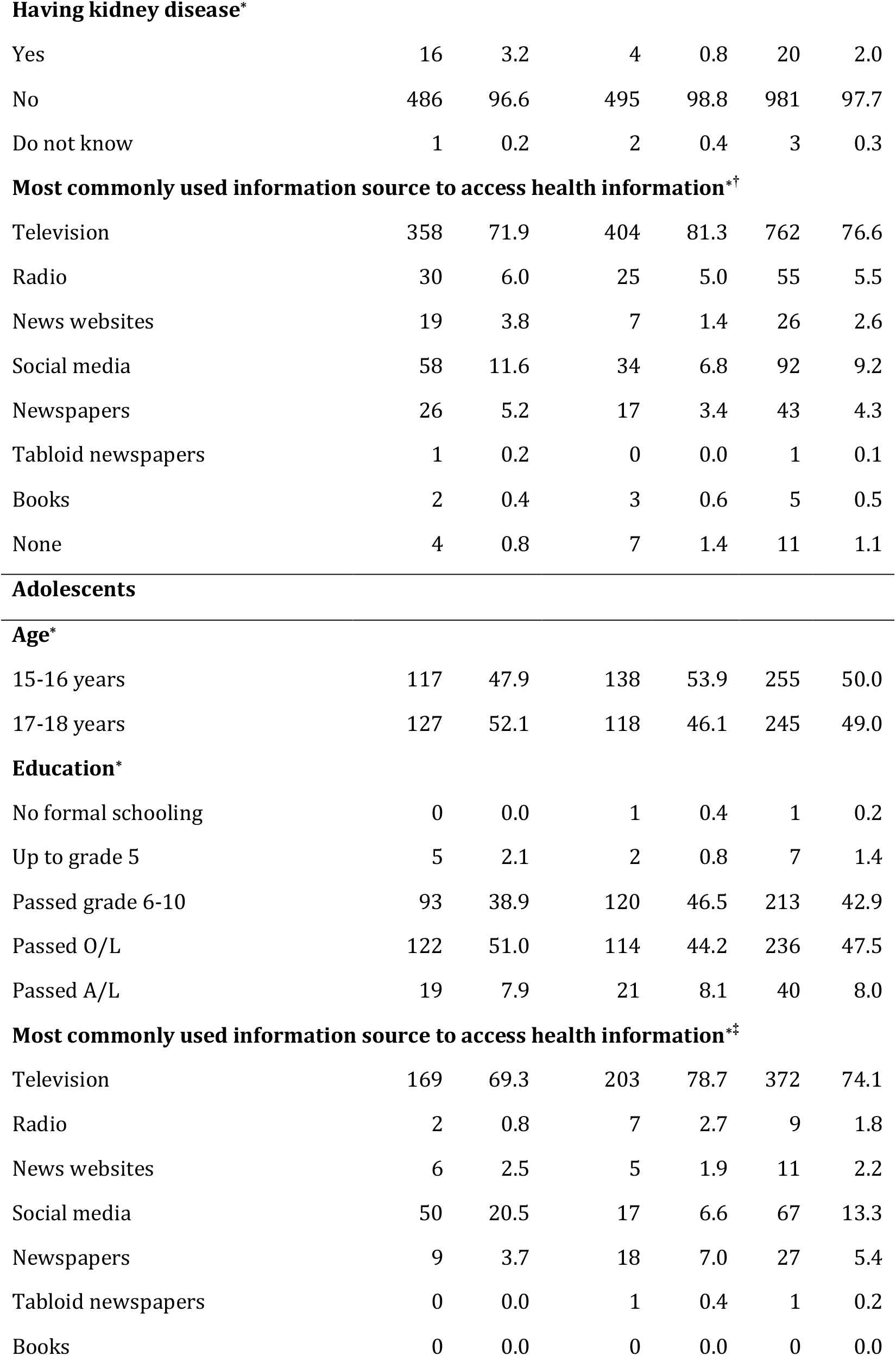
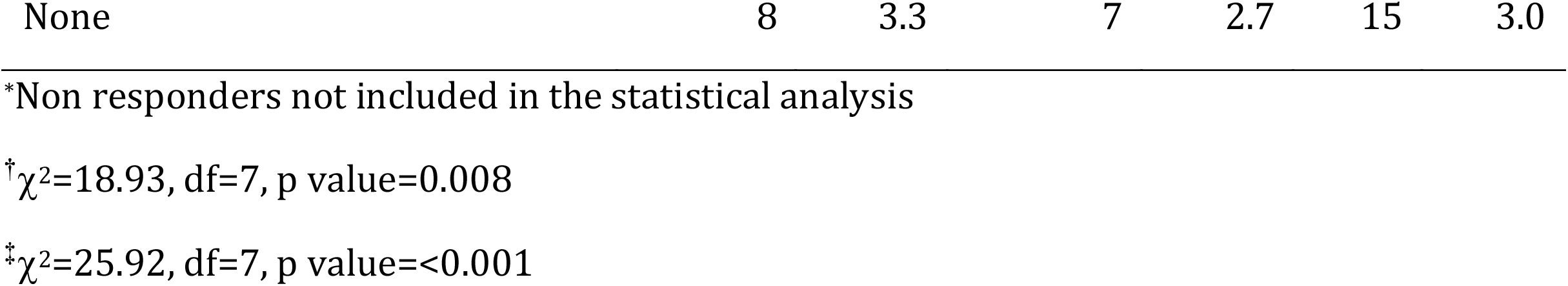
Sociodemographic characteristics of the adults and adolescents

|  | Male |  | Female |  | All |  |
| --- | --- | --- | --- | --- | --- | --- |
|  | n | % | n | % | n | % |
| **Adults** |  |  |  |  |  |  |
| **Age\*** |  |  |  |  |  |  |
| 19-25 | 43 | 8.5 | 32 | 6.4 | 75 | 7.4 |
| 26-40 | 137 | 27.1 | 167 | 33.3 | 304 | 30.2 |
| 41-55 | 156 | 30.9 | 176 | 35.1 | 332 | 33.0 |
| 56-70 | 135 | 26.7 | 110 | 21.9 | 245 | 24.3 |
| >70 | 34 | 6.7 | 17 | 3.4 | 51 | 5.1 |
| **Education\*** |  |  |  |  |  |  |
| No formal schooling | 12 | 2.4 | 8 | 1.7 | 20 | 2.1 |
| Up to grade 5 | 23 | 4.7 | 29 | 6.0 | 52 | 5.3 |
| Passed grade 6-10 | 82 | 16.7 | 73 | 15.1 | 155 | 15.9 |
| Passed O/L | 163 | 33.2 | 165 | 34.2 | 328 | 33.7 |
| Passed A/L | 155 | 31.6 | 161 | 33.4 | 316 | 32.5 |
| Degree and Above | 55 | 11.2 | 43 | 8.9 | 98 | 10.1 |
| no response | 1 | 0.2 | 3 | 0.6 | 4 | 0.4 |
| **Marital status\*** |  |  |  |  |  |  |
| Never Married | 67 | 13.2 | 46 | 9.1 | 113 | 11.1 |
| Married | 422 | 83.1 | 409 | 80.5 | 831 | 81.8 |
| Separated/Divorced/Widowed | 6 | 1.2 | 30 | 5.9 | 36 | 3.5 |
| no response | 13 | 2.6 | 23 | 4.5 | 36 | 3.5 |
| **Having hypertension\*** |  |  |  |  |  |  |
| Yes | 129 | 25.4 | 141 | 27.9 | 270 | 26.7 |
| No | 377 | 74.4 | 365 | 72.1 | 742 | 73.2 |
| Do not know | 1 | 0.2 | 0 | 0.0 | 1 | 0.1 |
| **Having Heart disease\*** |  |  |  |  |  |  |
| Yes | 49 | 9.7 | 28 | 5.6 | 77 | 7.6 |
| No | 455 | 90.3 | 475 | 94.4 | 930 | 92.4 |
| Do not know | 0 | 0.0 | 0 | 0.0 | 0 | 0.0 |
| **Having kidney disease\*** |  |  |  |  |  |  |
| Yes | 16 | 3.2 | 4 | 0.8 | 20 | 2.0 |
| No | 486 | 96.6 | 495 | 98.8 | 981 | 97.7 |
| Do not know | 1 | 0.2 | 2 | 0.4 | 3 | 0.3 |
| **Most commonly used information source to access health information\*†** |  |  |  |  |  |  |
| Television | 358 | 71.9 | 404 | 81.3 | 762 | 76.6 |
| Radio | 30 | 6.0 | 25 | 5.0 | 55 | 5.5 |
| News websites | 19 | 3.8 | 7 | 1.4 | 26 | 2.6 |
| Social media | 58 | 11.6 | 34 | 6.8 | 92 | 9.2 |
| Newspapers | 26 | 5.2 | 17 | 3.4 | 43 | 4.3 |
| Tabloid newspapers | 1 | 0.2 | 0 | 0.0 | 1 | 0.1 |
| Books | 2 | 0.4 | 3 | 0.6 | 5 | 0.5 |
| None | 4 | 0.8 | 7 | 1.4 | 11 | 1.1 |
| **Adolescents****Age\*** |  |  |  |  |  |  |
| 15-16 years | 117 | 47.9 | 138 | 53.9 | 255 | 50.0 |
| 17-18 years | 127 | 52.1 | 118 | 46.1 | 245 | 49.0 |
| **Education\*** |  |  |  |  |  |  |
| No formal schooling | 0 | 0.0 | 1 | 0.4 | 1 | 0.2 |
| Up to grade 5 | 5 | 2.1 | 2 | 0.8 | 7 | 1.4 |
| Passed grade 6-10 | 93 | 38.9 | 120 | 46.5 | 213 | 42.9 |
| Passed O/L | 122 | 51.0 | 114 | 44.2 | 236 | 47.5 |
| Passed A/L | 19 | 7.9 | 21 | 8.1 | 40 | 8.0 |
| **Most commonly used information source to access health information\*‡** |  |  |  |  |  |  |
| Television | 169 | 69.3 | 203 | 78.7 | 372 | 74.1 |
| Radio | 2 | 0.8 | 7 | 2.7 | 9 | 1.8 |
| News websites | 6 | 2.5 | 5 | 1.9 | 11 | 2.2 |
| Social media | 50 | 20.5 | 17 | 6.6 | 67 | 13.3 |
| Newspapers | 9 | 3.7 | 18 | 7.0 | 27 | 5.4 |
| Tabloid newspapers | 0 | 0.0 | 1 | 0.4 | 1 | 0.2 |
| Books | 0 | 0.0 | 0 | 0.0 | 0 | 0.0 |
| None | 8 | 3.3 | 7 | 2.7 | 15 | 3.0 |

\*Non responders not included in the statistical analysis
<sup>†</sup> $\chi^2=18.93$ , df=7, p value=0.008
<sup>‡</sup> $\chi^2=25.92$ , df=7, p value=<0.001

Taking into account all the 1016 household of adults included in the survey, the median house hold size was estimated as four individuals (IQR: 2-5 individuals).

A total of 505 adolescents participated in this study (Table 1). The median age of the adolescents was 16 years (IQR: 15-18 years) and the majority were Sinhalese (n=411; 82.5%). Though both male and female adolescents most commonly used TV for health information (n=372; 74.1%), the proportion of female adolescents (n=203; 78.7%) who did so was significantly higher than the male adolescents (n=169; 69.3%) (p<0.001). One important finding was that only 9.2% of the adults and 13.3% of the adolescents used online sources for information, which was a surprising finding considering the fact that the internet penetration of the country is 34%^20^.

### Salt and health: knowledge of adults and adolescents

As indicated in Table 2, only 40.1% (n=407) of adults and only 200 (39.6%) of adolescents knew the recommended limit of salt per person as 5g/day or less. A significantly higher proportion of adult females (n=220; 43.3%) knew the correct daily allowance of salt for adults, compared to adult males (n=187; 36.8%) (p=0.032). Among adolescents, the proportion who were aware of the recommended limit of salt per person as 5g/day or less was similar among males (n=97; 39.6% and females (n=103; 39.6%) (p=0.996). Exploring knowledge about adverse health effects of high salt consumption, the study revealed that high proportions of adults (n=846; 83.3%) and adolescents (n=354; 70.1%) knew that high salt intake is a risk factor for hypertension. Contrastingly, only less than half adults (n=413; 40.6%) and adolescents (n=173; 34.3%) knew that high salt intake is a risk factor for kidney disease. The proportion adults (n=229; 25.5%) and adolescents (n=212; 26.4%) who knew that high salt intake is a risk factor for stomach cancer was even less. There were no statistically significant differences of knowledge on adverse health effects of high salt consumption between males and females among either adults or adolescents.

**Table 2.**
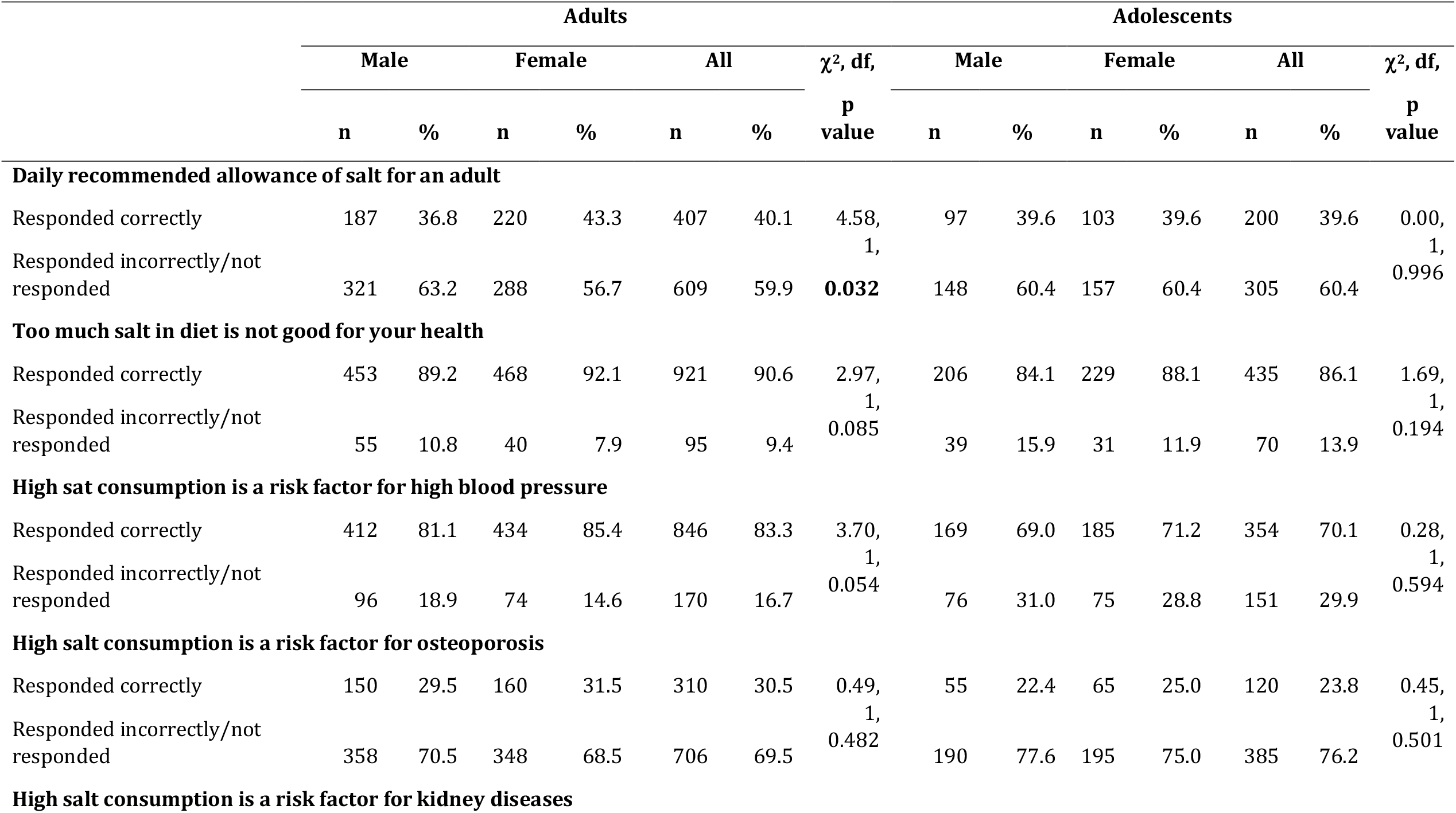
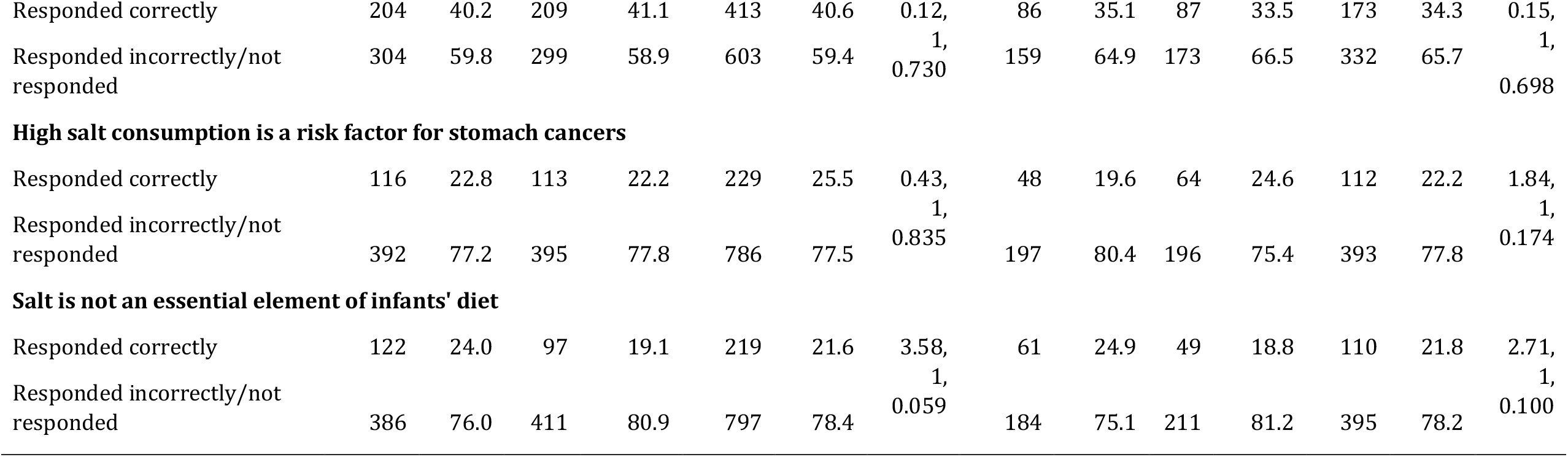
Knowledge on salt and health among adolescents and adults

### Salt and health: attitudes of adults and adolescents

As illustrated in Table 3, approximately half of the adults (n=488; 48.3%) (males- n=243; 48.1%: female- n=245; 48.5%) and adolescents (n=231; 45.7%) (males- n=110; 44.9%: female- n=121; 46.9%) believed that they are eating ‘just the right amount of salt’. This belief of eating just the right amount of salt was not significantly different between males and females (adults- p=0.758, adolescents- p=0.889).

**Table 3.** Attitudes/opinion on salt and health among adolescents and adults

|  | Adults |  |  |  |  |  |  | Adolescents |  |  |  |  |  |  |
| --- | --- | --- | --- | --- | --- | --- | --- | --- | --- | --- | --- | --- | --- | --- |
| | Male | | Female | | All | | $\chi^2$, df, | Male | | Female | | All | | $\chi^2$, df, |
|  | n | % | n | % | n | % | p value | n | % | n | % | n | % | p value |
| **Opinion about amount of salt or salty food they consumed** |  |  |  |  |  |  |  |  |  |  |  |  |  |  |
| Far too much | 20 | 4.0 | 27 | 5.3 | 47 | 4.7 | 1.88, 4,<br>0.758 | 11 | 4.5 | 8 | 3.1 | 19 | 3.8 | 1.13, 4,<br>0.889 |
| Too much | 148 | 29.3 | 159 | 31.5 | 307 | 30.4 |  | 77 | 31.4 | 76 | 29.2 | 153 | 30.3 |  |
| Just the right<br>amount | 243 | 48.1 | 245 | 48.5 | 488 | 48.3 |  | 110 | 44.9 | 121 | 46.5 | 231 | 45.7 |  |
| Too little | 44 | 8.7 | 46 | 9.1 | 90 | 8.9 |  | 11 | 4.5 | 12 | 4.6 | 23 | 4.6 |  |
| Far too little | 13 | 2.6 | 9 | 1.8 | 22 | 2.2 |  | 3 | 1.2 | 2 | 0.8 | 5 | 1.0 |  |
| No comment* | 37 | 7.3 | 19 | 3.8 | 56 | 5.5 |  | 33 | 13.5 | 41 | 15.8 | 74 | 14.6 |  |
| **Opinion on importance of lowering own dietary salt** |  |  |  |  |  |  |  |  |  |  |  |  |  |  |
| Very<br>important | 265 | 52.2 | 295 | 58.2 | 560 | 55.2 | 2.10, 2, 0.<br>0.350 | 88 | 35.9 | 111 | 42.7 | 199 | 39.4 | 9.31, 2,<br>**0.010** |
| Somewhat<br>important | 186 | 36.6 | 177 | 34.9 | 363 | 35.8 |  | 110 | 44.9 | 111 | 42.7 | 221 | 43.8 |  |
| Not at all<br>important | 26 | 5.1 | 21 | 4.1 | 47 | 4.6 |  | 24 | 9.8 | 9 | 3.5 | 33 | 6.5 |  |
| No comment* | 31 | 6.1 | 15 | 2.8 | 46 | 4.4 |  | 23 | 9.4 | 29 | 11.2 | 52 | 10.3 |  |
| **Opinion on how easy to reduce the salt consumption** |  |  |  |  |  |  |  |  |  |  |  |  |  |  |
| Very easy | 71 | 14.0 | 75 | 14.8 | 146 | 14.4 | 7.12, 5,<br>0.212 | 28 | 11.4 | 23 | 8.8 | 51 | 10.1 | 7.50, 5,<br>0.186 |
| Easy | 301 | 59.3 | 284 | 56.0 | 585 | 57.6 |  | 128 | 52.2 | 167 | 64.2 | 295 | 58.4 |  |
| Not difficult | 60 | 11.8 | 86 | 17.0 | 146 | 14.4 |  | 45 | 18.4 | 33 | 12.7 | 78 | 15.4 |  |
| Not easy | 34 | 6.7 | 27 | 5.3 | 61 | 6.0 |  | 15 | 6.1 | 13 | 5.0 | 28 | 5.5 |  |
| Difficult | 37 | 7.3 | 29 | 5.7 | 66 | 6.5 |  | 24 | 9.8 | 20 | 7.7 | 44 | 8.7 |  |
| Very difficult | 4 | 0.8 | 5 | 1.0 | 9 | 0.9 |  | 3 | 1.2 | 3 | 1.2 | 6 | 1.2 |  |
| No comment* | 1 | 0.2 | 2 | 0.2 | 3 | 0.2 |  | 2 | 0.8 | 1 | 0.4 | 3 | 0.6 |  |

\*No comments category was not included in the analysis

Of all the participants, 55.2% (n=560) (males- n=265; 52.2%: female-n= 295; 58.2%) of the adults and 39.4% (n=199) (males- n=88; 35.9%: female- n=111; 42.7%) of the adolescents opined that it is very important to lower their own dietary salt consumption. This pattern of attitude was not significantly different between adult males and females (p=0.350) but showed a significant difference between adolescent males and females (p=0.010) with more females attributing importance to reducing salt consumption.

When inquired into the participants’ attitude on how easy it will be for the respondent to reduce the salt consumption, a majority of the adults (n=731, 72.%, males-n=372, 73.3%: female- n=359,70.8%) and adolescents (n=346, 68.50% (males-n=156, 63.3%: female n=190,73%) believed that it is very easy/easy to reduce their dietary salt consumption. There were no statistically significant differences of this attitude between males and females among either adults (p=0.212) or adolescents (p=0.186) (Table 3).

### Salt and health: cooking practices of adults

Inquiries into type of salt from adults indicate the choices powered and crystal with approximately half (n=456, 47%) choosing either. In Sri Lanka, salt does not come in dispensable packs and most (n=683, 69.5%) stored salt in a bottle/container. As illustrated in Table 4, the study attempted to also indirectly quantify the salt consumption of the adults by inquiring the adults into the amount of monthly purchases of salt to the households and the number of household members. The median total monthly household salt purchased was 1100g (IQR 700-1450g) with the median house hold size estimated as 4 individuals (IQR: 2-5 individuals). Approximate estimates indicated that per capita consumption of salt to be nearly double the recommended amount among the household members of the adult respondents of the present study.

**Table 4.** Cooking practices related to salt among adults

| | Male | | Female | | All | | $\chi^2$, df,<br>p value* |
| --- | --- | --- | --- | --- | --- | --- | --- |
|  | N | % | n | % | n | % |  |
| Household cooking practices |  |  |  |  |  |  |  |
| Type of salt usually purchased |  |  |  |  |  |  |  |
| Always powdered salt | 189 | 39.9 | 183 | 36.8 | 372 | 38.3 | - |
| Always crystal salt | 65 | 13.7 | 78 | 15.7 | 143 | 14.7 |  |
| No preference | 220 | 46.4 | 236 | 47.5 | 456 | 47.0 |  |
| Amount of salt purchased per month (g) | Median | IQR | Median | IQR | Median | IQR |  |
| Powdered | 500 | 200-1000 | 500 | 150-1000 | 500 | 200-1000 | - |
| Crystal | 400 | 250-800 | 400 | 225-500 | 400 | 250-800 |  |
| Total | 1100 | 700-1450 | 1000 | 625-1450 | 1200 | 800-1600 |  |
| How salt is stored at home |  |  |  |  |  |  |  |
| In the packet itself | 52 | 10.8 | 57 | 11.4 | 109 | 11.1 | - |
| In a bottle/container | 334 | 69.2 | 349 | 69.8 | 683 | 69.5 |  |
| Dissolved in a coconut shell/bottle | 48 | 9.9 | 49 | 9.8 | 97 | 9.9 |  |
| Do not know | 49 | 10.1 | 45 | 9.0 | 94 | 9.6 |  |
| Personal cooking practices |  |  |  |  |  |  |  |
| Frequency of cooking meals at home |  |  |  |  |  |  |  |
| Always | 81 | 15.9 | 420 | 82.7 | 501 | 49.4 |  |
| Most of the time | 33 | 6.5 | 43 | 8.5 | 76 | 7.5 | 526.65, 4,<br><0.001 |
| Sometimes | 74 | 14.6 | 26 | 5.1 | 100 | 9.8 |  |
| Rarely | 77 | 15.2 | 7 | 1.4 | 84 | 8.3 |  |
| Never | 243 | 47.8 | 12 | 2.4 | 255 | 25.1 |  |
| **Adding salt while cooking** | (n=265) |  | (n=496) |  | (n=761) |  |  |
| Yes | 230 | 86.8 | 424 | 85.5 | 654 | 85.9 | 0.66, 1, |
| No | 35 | 13.2 | 72 | 14.5 | 107 | 14.1 | 0.418 |
| **Adding salt to rice while cooking** | (n=232) |  | (n=424) |  | (n=656) |  |  |
| Yes | 116 | 50.0 | 230 | 54.2 | 346 | 52.7 | 1.04, 1, |
| No | 116 | 40.0 | 194 | 45.8 | 310 | 47.3 | 0.309 |
| **How salt is added while cooking** | (n=232) |  | (n=424) |  | (n=656) |  |  |
| Add the amount I think is needed at once | 113 | 48.7 | 199 | 46.9 | 312 | 47.6 | 2.88, 3, |
| Add little at the beginning and keep adding | 114 | 49.1 | 205 | 48.3 | 319 | 48.6 | 0.410 |
| Add required amount at the end | 5 | 2.2 | 19 | 4.5 | 24 | 3.7 |  |
| Other | 0 | 0.0 | 1 | 0.2 | 1 | 0.2 |  |
| **Use of seasoning cubes while cooking at home** | (n=232) |  | (n=424) |  | (n=656) |  |  |
| Yes | 82 | 35.3 | 159 | 37.5 | 241 | 36.7 | 0.30, 1, |
| No | 150 | 64.7 | 265 | 62.5 | 415 | 63.3 | 0.584 |
| **Frequency of adding seasoning cubes** | (n=82) |  | (n=159) |  | (n=241) |  |  |
| Always | 2 | 2.4 | 5 | 3.1 | 7 | 2.9 | 3.10, 2, |
| Sometimes | 49 | 59.8 | 76 | 47.8 | 125 | 51.9 | 0.212 |
| Rarely | 31 | 37.8 | 78 | 49.1 | 109 | 45.2 |  |
| **Use of soy sauce while cooking at home** | (n=232) |  | (n=424) |  | (n=656) |  |  |
| Yes | 50 | 21.6 | 90 | 21.2 | 140 | 21.3 | 0.01, 1, |
| No | 182 | 78.4 | 334 | 78.8 | 516 | 78.7 | 0.922 |

Exploring cooking practices revealed that among adults, a significantly higher proportion of females (n=420; 83.0%) ‘always’ cooked meals at home compared to males (n=81; 15.9%) (p=<0.001). Approximately half who reported cooking, (n=346, 52.7%) (males- n=116; 50.2%: female-n= 230; 54.4%) said that they add salt to rice and almost all (n=631, 96.2%) (males- n=227, 97.8%: female- n=404; 95.2%) reported that they add salt prior to or while cooking food. Females and males did not show a significant difference in these practices (Table 4).

### Salt and health: dietary practices of adults and adolescents

As shown in Table 5, regarding the habit of adding salt to cooked food at the table, of the adults, only 16.7% (n=169) (males- n=88; 17.4%: female- n=81; 16.0%) and of the adolescents, only 13.7%(n=68) (males- n=26; 10.7%: n=42; female-16.6%) reported that they add salt while eating food already cooked with salt. The practice of daily ‘eating out’ was quite rare among adults though was higher among males (breakfast n=; 3.6% (males- n=28; 5.5%; females-n=9; 1.8% p=<0.001), lunch (n=34; 3.3% (males- n=27; 5.3%; females-n=7; 1.4% p=<0.001%) and dinner (n=19; 1.9% (males- n=15; 3.0%; females- n=4; 0.8% p=0.025%). The frequency of eating lunch and dinner out of the home were significantly different among male and female adolescents (lunch p=<0.001; dinner p=0.045) with higher proportions of males consuming these more frequently. Almost all the adults (n=971; 95.6% (males- n=483; 95.1%; females- n=488; 96.1%) and adolescents (n=489, 96.8% (males- n=238; 97.1%; females- n=251; 96.5%) reported that they have never ordered food online and there was no significant difference between males and females (adults-p=0.345; adolescents- p=0.812).

**Table 5.**
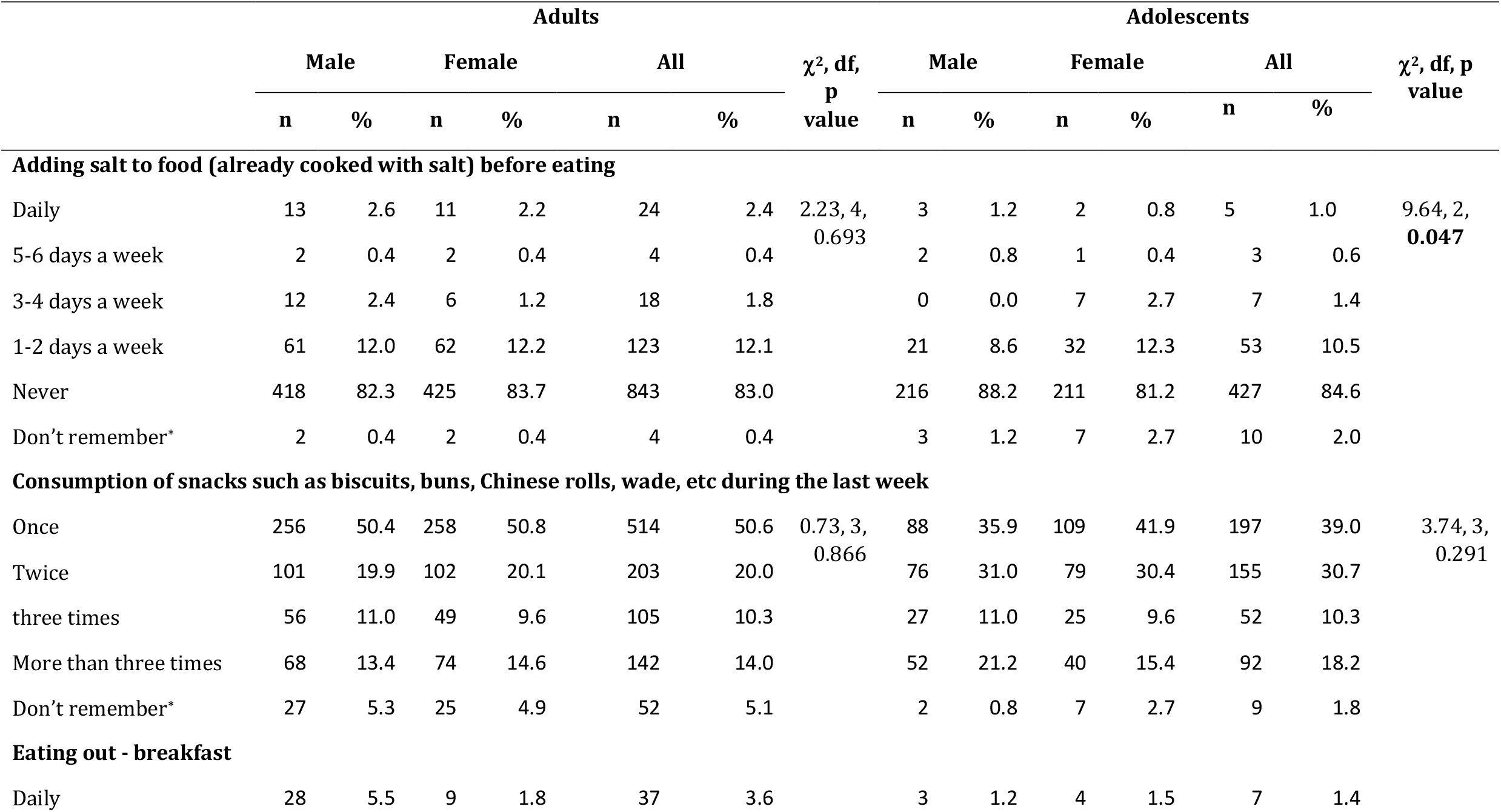
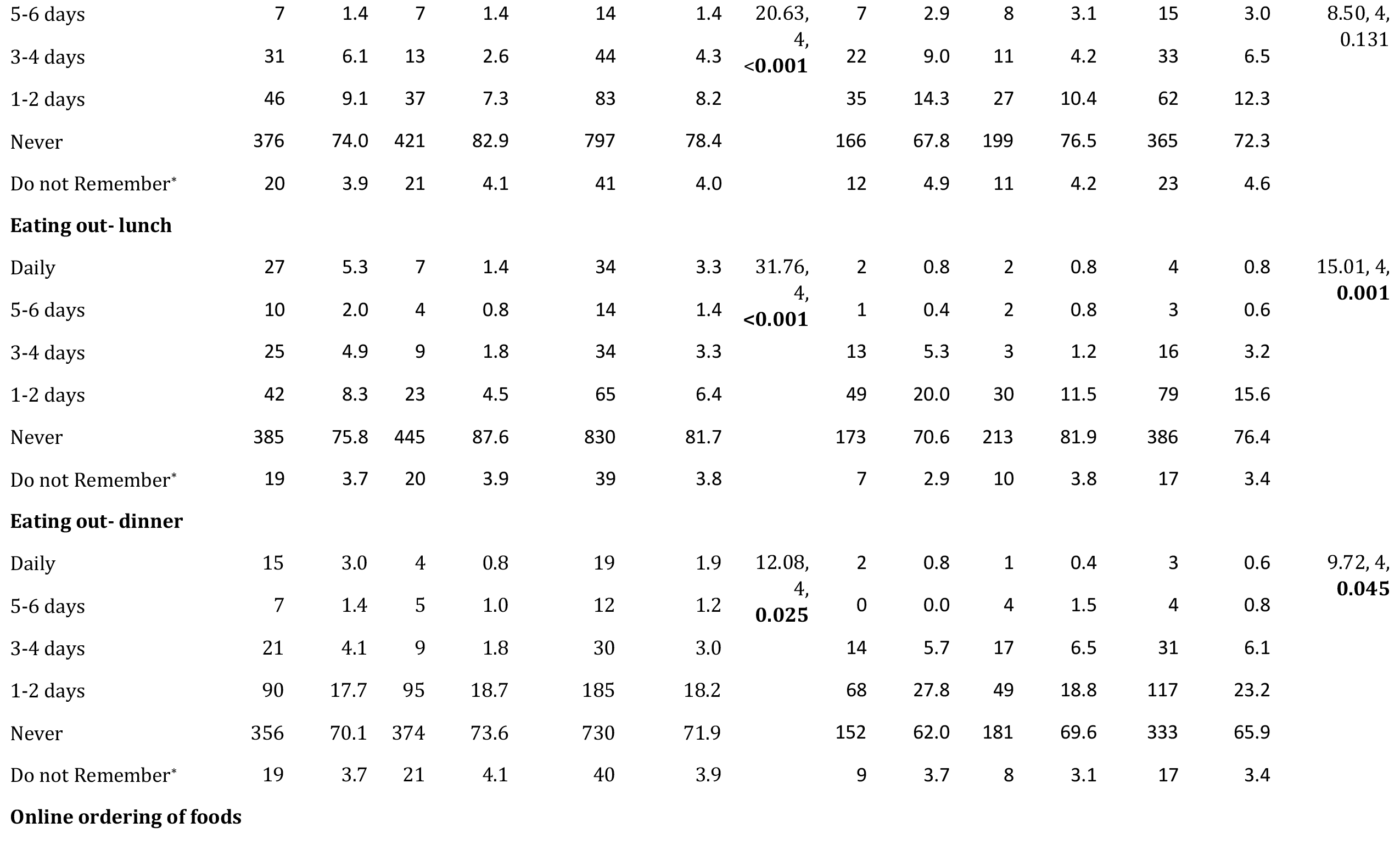
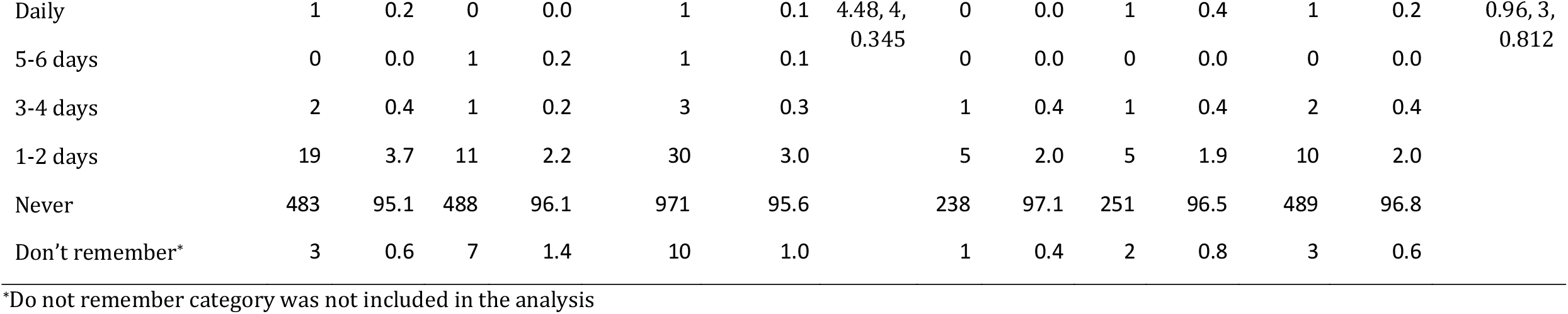
Dietary practices in the week prior to the survey among adolescents and adults

## Discussion

The present study generated important evidence to inform the designing of communication material in implementing an effective behavior change communication campaign of the NSRS 2018-2022. The results of this study indicated that majority of Sri Lankan adults (59.9%) and adolescents (60.4%) were not adequately aware of the recommended daily salt consumption. This could be a main deterrent and a crucial knowledge gap that affect Sri Lanka’s efforts to implement the NSRS’s aim to reduce population level dietary salt consumption. It was also noted that this finding is compatible with findings of the studies in other LMICs^14,16–18,21^. Furthermore, the indirect estimation of daily dietary salt consumption in our study showed that Sri Lankans consume higher than the daily requirement of salt.

These finding taken together with the finding that approximately half of the adults (48.3%) and adolescents (45.9%) believing that they are eating just the right amount of salt is very disturbing and reiterates the importance of the NSRS to implement an effective behavior change communication campaign. The findings of a vast majority of adults and adolescents knowing that high salt consumption is not good for health and its association with hypertension and that a majority (>55%) of adults and adolescents believing that it is very important to reduce their salt consumption can be considered as clues towards success. The finding of the present study related to 75% of adults and adolescents use television as their main source of health information should inform the NSRS of how to design the communication campaign ^22^. Because the use of social media was only 9.2% and 13.3% among adults and adolescents respectively, social media campaigns are unlikely to reach majority of population in Sri Lanka. In a 2019 global digital overview, it was stated that Asia’s social media landscape is different from the that of West, and only 24% of the south Asians use social media landscapes. Our finding confirms that local information platforms dominate the online platforms in Asian countries^20^.

Decreasing the exposure to salty tasting foods during early infancy is recommended^23^. Only around 22% of adults and adolescents knew that salt is not an essential element of infants’ diet. The well-established maternal and child health programs in Sri Lanka can be effectively utilized to initiate a long-term positive effect to the whole community and create a generation that is used to low salt diets.

With regards to practices to be addressed through the communication campaign, the study indicated that more than a half of the participants reported that they add salt to rice, which is a finding reported in previous studies as well indicating ineffective efforts to reduce salt consumption in the past years in Sri Lanka^8^. Rice is the main dish of Sri Lankan food and is taken in high quantities. Moreover, Sri Lankans mainly consume homemade food rather than packaged foods. Therefore, discouraging the practice of adding salt to rice could be an important intervention to reduce the salt consumption. Because home cooked food is prepared mostly by females and salt or seasoning cubes are added to food while being cooked, this highlights the need to target adult females in the communication campaign on salt reduction. This pattern was seen in a study conducted in Nepal as well, possibly because of similar socio-cultural backgrounds^14^.

A strength of our study is that this is the first island wide survey to assess knowledge, attitudes, and practices of dietary salt consumption in Sri Lanka. Though much care was taken to assess the self-reported practices related to cooking and diet in an objective manner, the limitation of recall bias cannot be ruled out. Another limitation is that the present study quantified the salt consumption of the study units indirectly by inquiring into monthly family household salt consumption and this can only be considered a proxy indicator.

In conclusion, the study identified gaps as well strengths in knowledge, attitudes and practices of Sri Lankans related to salt and health that should be addressed through the planned communication campaign of NSRS. Not being aware of the recommended dietary salt intake, practice of adding salt to rice and consumed much higher amounts of salt than that is recommended, while believing that they consume the right amount are some key gaps. Knowing the association of salt and health, agreeing that personal dietary salt reduction as important and mostly eating home cooked food are the strengths on which the communication campaign can lever on. The survey identified adult females to be a key target group for the campaign and television is recommended as the mode of delivery.

## Data Availability

Data available with the corresponding author

## Acknowledgements

The support of staff and students of Department of Health promotion of the Rajarata University of Sri Lanka and Dr Padmal de Silva of Word Health Organization Country Office for Sri Lanka are acknowledged.

## Author contribution

AJ, NG, AS, AW, CW, AA contributed to design the study and AJ, NG, PR carried out the study and analysed the data and AJ, NG, AS interpreted the findings and wrote the first draft of the article and all authors provided critical revisions of the draft and all authors read and approved the final manuscript.

